# Testing of retail cheese, butter, ice cream and other dairy products for highly pathogenic avian influenza in the US

**DOI:** 10.1101/2024.08.11.24311811

**Authors:** David L. Suarez, Iryna V. Goraichuk, Lindsay Killmaster, Erica Spackman, Nicole J. Clausen, Tristan J. Colonius, Cynthia L. Leonard, Monica L. Metz

## Abstract

The recent outbreak of highly pathogenic avian influenza (HPAI) in dairy cows has created public health concerns about the potential of consumers being exposed to live virus from commercial dairy products. Previous studies support that pasteurization effectively inactivates avian influenza in milk and an earlier retail milk survey showed viral RNA, but no live virus could be detected in the dairy products tested. Because of the variety of products and processing methods in which milk is used, additional product testing was conducted to determine if HPAI viral RNA could be detected in retail dairy samples, and for positive quantitative real-time RT-PCR (qRT-PCR) samples to be tested for presence of live virus. Revised protocols were developed to extract RNA from solid dairy products including cheese and butter. The solid dairy product was mechanically liquified with garnet and zirconium beads in a bead beater diluted 1 to 4 with BHI media. This pre-processing step was suitable in allowing efficient RNA extraction with standard methods. Trial studies were conducted with different cheese types with spiked in avian influenza virus to show that inoculation of the liquified cheese into embryonating chicken eggs was not toxic to the embryos and allowed virus replication. A total of 167 retail dairy samples, including a variety of cheeses, butter, ice cream, and fluid milk were collected as part of nationwide survey. A total of 17.4% (29/167) of the samples had detectable viral RNA by qRT-PCR targeting the matrix gene, but all samples were negative for live virus after testing with embryonating egg inoculation. The viral RNA was also evaluated by sequencing part of the hemagglutinin gene using a revised protocol optimized to deal with the fragmented viral RNA. The sequence analysis showed all viral RNA positive samples were highly similar to previously reported HPAI dairy cow isolates. Using the revised protocols, it was determined that HPAI viral RNA could be detected in a variety of dairy products, but existing pasteurizations methods effectively inactivate virus assuring consumer safety.

## Introduction

Highly pathogenic avian influenza (HPAI) has emerged as a global animal health threat for poultry, wild birds, and marine mammals(Graziosi et al., 2024; Swayne, Suarez, & Sims, 2020). Both H5 and H7 HPAI outbreaks have been reported worldwide in the past 2 years, but the biggest concern is the H5 goose/Guangdong lineage, which was first identified in 1996. This lineage of virus was originally identified in poultry, but the virus appeared to spill back into wild birds in the early 2000s, and has since become endemic in the wild bird population (Lee et al., 2017). Despite intensive control measures in poultry, which have included depopulation of infected flocks, vaccination in some countries, enhanced biosecurity, and quarantine and movement control programs, the temporary successes in control have been undermined by the constant pressure of reintroduction from the endemic virus in wild birds (Swayne, Suarez, & Sims, 2020; Youk et al., 2023).

Highly pathogenic avian influenza virus is a segmented RNA virus in the Orthomyxyoviridae family and Alphainfluenzavirus genus or more commonly referred to as type A influenza virus. Type A influenza viruses include many unique lineages of virus that replicate and transmit easily within a specific host population and human, swine, equine, avian, and canine lineages are well described(Suarez, 2016). However, the inter-species barrier for influenza A viruses is low and it is common for viruses to jump to new species, although sustained transmission is rare(Yamaji et al., 2020). Influenza A viruses are segmented, and reassortment of gene segments is commonly observed when two unique influenza A viruses concurrently infect the same host. The other salient feature of influenza A virus is the plasticity of the hemagglutinin protein, which can change its antigenic properties to avoid the host immune response. The ability of influenza viruses to rapidly change and adapt to new species is commonly called drift and shift(Suarez, 2016). The highly pathogenic avian influenza virus currently circulating in the U.S. is evidence of this shift and drift(Youk et al., 2023).

The goose/Guangdong lineage, first identified in 1996, has constantly reassorted with many other influenza viruses in the last 28 years, and the only constant has been the hemagglutinin gene. The hemagglutinin gene has antigenically drifted extensively since 1996 and a complex clade system was developed to track these antigenic changes(“Revised and updated nomenclature for highly pathogenic avian influenza A (H5N1) viruses,” 2014). The dominant lineage for the current outbreak in North America, South America and Europe is clade 2.3.4.4b. The first detection of clade 2.3.4.4b in the Americas was in Canada in late 2021(Caliendo et al., 2022). The virus was most closely related to viruses circulating in Europe and the neuraminidase and all 6 internal genes were of Eurasian lineage. This Eurasian origin virus began to reassort with low pathogenic avian influenza viruses in the Americas creating different genotypes of virus with different internal gene combinations. A genotype system was developed to track the viruses with the different gene combinations, and some combinations have been associated with widespread poultry and wild bird detections(Youk et al., 2023) Although there are unique viruses circulating in the Americas, the phenotype of the virus has not appeared to change with high mortality in chickens and turkeys and variable mortality in other species.

In March of 2024 HPAI was detected in dairy cattle in the U.S. and sequence analysis showed it to be a clade 2.3.4.4b virus with the previously uncommon B3.13 genotype, which has 4 genes of North American origin (NS, PB1, PB2 and NP) and 4 genes of Eurasian origin (HA, NA, MA, and PA) (Burrough et al., 2024; Caserta et al., 2024). With the single introduction into dairy cows, the virus has spread into at least 178 dairy farms in 13 different states as of early August 2024 ( HPAI Confirmed Cases in Livestock | Animal and Plant Health Inspection Service (usda.gov)). The viral infection primarily has caused mastitis with a decrease both in milk quality and milk volume in affected cows. Some cows have had more severe disease and require supportive care. The mastitis is usually self-limiting and milk quality and volume will recover, although cows might not return to pre-infection levels of production. The infection is primarily in the mammary gland and high levels of infectious virus can be found in unpasteurized milk. The infected cow’s immune response will produce antibody that correlates to a rapid decline in detectable viable virus, although viral RNA could still be detected for several weeks (Caserta et al., 2024). The presence of HPAI in unpasteurized milk creates the potential for zoonotic spread to humans through consumption of the virus in milk and other dairy products.

Influenza A viruses are sensitive to heat and studies have shown that high temperature/short time continuous flow pasteurization, which is the industry standard, effectively inactivates the virus in the milk (Spackman, submitted). The effectiveness of current pasteurization methods was further demonstrated by a survey of retail milk and a small number of other dairy products that showed the detection of viral RNA in some dairy samples, but no viable virus was detected by attempting to culture the virus in embryonating chicken eggs, which is a sensitive detection method (Spackman et al., 2024).

According to the U.S. Code of Federal Regulation cheeses must be made from pasteurized milk unless there is an alternative in the standard of identity ([eCFR :: 21 CFR Part 133 -- Cheeses and Related Cheese Products](https://www.ecfr.gov/current/title-21/chapter-I/subchapter-B/part-133)) to use raw milk in combination with aging the cheese at a temperature of not less than 35 °F for at least 60 days. Pasteurized milk cheeses are those made from milk that has been heated to minimum temperatures and times found in 21 CFR 133.3(d) (https://www.ecfr.gov/current/title-21/chapter-I/subchapter-B/part-133/subpart-A/section-133.3), for example the milk is treated to 161 °F for 15 seconds. Raw milk cheeses use milk that either has not been heat-treated or has been heat-treated to a temperature/time combination that is less than pasteurization parameters which may be referred to as thermization.

The cheese making process varies depending on the type of cheese being made. Cheddar cheese may be made with either pasteurized milk or raw milk combined with aging according to 21 CFR 133.113 (eCFR :: 21 CFR 133.113 -- Cheddar cheese.). According to McSweeney, 2007, cheddar cheese is made with mesophilic lactic acid bacteria. Milk is heated to around 86 °F allowing the culture to grow, and rennet is then added. The curds and whey are then heated to 98.6 °F - 102.2 °F long enough to achieve about a 5.4 pH. The cheese curd would not be heated further in the manufacturing process of cheddar cheese(McSweeney, 2007).

For mozzarella, thermophilic lactic acid bacteria are used. Pasteurized milk is heated to around 88 °F (21 CFR 133.155) allowing the culture to grow and rennet is then added. Once curd is formed, the curd and whey are heated to about 104°F -109.4 °F. When the desired pH is achieved, milled curd is immersed in hot water ranging from 149°F -176 °F and then mechanically molded by a rotating single-screw cooker/stretcher where the curd is kept at a temperature of 122°F-131°F. Mozzarella cheese would not undergo any further heating processes(Minervini, Costantino, & De Angelis, 2022).

Pasteurized processed cheeses can be made from both pasteurized milk cheeses and raw milk cheeses. The cheeses are ground/milled and mixed with the aid of heat according to 21 CFR 133.169 (eCFR :: 21 CFR 133.169 -- Pasteurized process cheese.). The cheese mixture is heated for not less than 30 seconds at a temperature of not less than 150 °F. The process cheese would not undergo any further heating process.

In a continuation of the original retail dairy product survey, additional testing was conducted to evaluate other types of dairy products, including cheese, ice cream, and butter. Most of these products are prepared from pasteurized milk, and the risk of detecting viable virus was low, but multiple raw milk cheeses were also tested. This study looks to build on previous work to develop robust methods for testing different types of retail milk products for the presence of viral RNA and if present determine if viable virus remains in the sample.

## METHODS

### Sample Collection

The U.S. Food and Drug Administration (FDA) contracted with a third party to collect and single blind 167 samples of milk and dairy products from retail locations throughout the United States that were sent to the U.S. Department of Agriculture (USDA) Agricultural Research Service (ARS) for analysis between June 18 – July 31, 2024. In this study, FDA expanded upon a previous retail dairy sampling survey to include pasteurized fluid milk in underrepresented geographic areas from the initial study and included dairy product types not analyzed in the previous survey including pasteurized milk products including cheeses, butter, ice cream, and unpasteurized or raw milk aged cheese and sample origin focused on regions with herds confirmed to have H5N1 HPAI virus infections.

A sample submission process using single blinding was developed to survey the market in response to input from industry and state regulatory partners that had concerns that identifying specific brands or processors in this study could have unintended consequences on the milk supply. The study was designed to remove identifying marks such as brand names and lot codes by aseptically repacking the sample and sending it to USDA ARS without any information about the sample origin. The study design retained meta-data on the state of processing and product types for each product. While meta-data on the state of processing has value to state regulators, milk can be produced in one or more states, processed in another, and sold in yet a third state. If analytical results had identified viable virus, information about the product, including manufacturer information for the related sample, could be unblinded so FDA could take appropriate public health action. Controls were in place to ensure the products maintained their appropriate temperatures (i.e., <7°C) throughout the collection and shipping processes. Samples were retained in the event a sample needed re-analysis.

Fluid milk samples consisted of different fat contents and included whole, 2%, 1%, skim, and heavy cream. Eighty pasteurized fluid milk samples were collected representing processors from 27 states (AL, AZ, CA, CT, DE, ID, IL, KY, LA, MD, ME, MS, MT, NC, NC, NE, NH, NM, NJ, OH, PA, RI, SD, TN, VA, VT, WI). Sampling was designed to be representative based on pounds of milk used in production per state with sample numbers also allocated in consideration of the previous FDA fluid milk survey to fill remaining geographic gaps with a minimum of three samples per state.

Cheeses included in the study were cheddar cheese, processed cheese, cream cheese, and mozzarella cheese made from pasteurized milk. Additionally, sampling was conducted on aged raw milk aged cheeses. All cheeses were without flavors or added ingredients, such as herbs. The cheese type was selected using data from USDA’s Economic Research Service on domestic per capita consumption. In total we sampled: 14 cheddar cheese samples representing processers in 9 states (CO, ID, MI, MN, NC, NM, OH, SD, TX); 12 mozzarella cheese representing processers in 8 states (CO, ID, MI, MN, OH, RI, SD, TX); 6 processed cheese samples representing processers in 4 states (ID, MI, OH, TX); 3 cream cheese samples representing processors in 2 states (MI, TX); 23 aged raw milk aged cheese samples representing processers in 5 states (ID, MN, NC, OH, TX). Additional dairy products analyzed included: 7 Butter samples representing processors across 5 states (OH, ID, MI, MN, NC); 22 vanilla ice cream samples were collected representing processers in 7 states (CO, ID, MI, NM, OH, SD, TX).

### Validation of Sample processing and Liquification

A variety of cheese samples (raw, pasteurized, hard, soft) were selected from local retailers to conduct preliminary trials to optimize extraction, PCR and egg inoculation procedures. Approximately 250 mg of each cheese sample was measured and added to either 0.5 ml or 1 ml brain heart infusion broth and 30 µl antibiotics (initial concentration penicillin G, 10,000 IU/ml; gentamicin, 1 mg/ml; kanamycin, 0.65 mg/ml; streptomycin, 2 mg/ml; and amphotericin B, 20 µg/ml).

Methods for liquifying cheese samples included: grinding with a pestle in a 2 ml vial; mincing with scalpel and vortexing; expelling through a 3-ml syringe (no needle); pressing through a fine wire mesh filter; grinding in 1.0 mm zirconium bead or in 500 µm garnet and 6mm zirconium satellite pre-filled vials (OPS Diagnostics, Lebanon, NJ) using a bead beater (FastPrep 24 -Classic, MP Biomedicals)(Das et al., 2008). Samples were liquified in the bead beater at 6.5 meters/second for 60s. Based on preliminary results where the product reached a liquid consistency, the remaining validation procedures were performed using 250 mg of sample in 0.5 ml BHI or 250mg sample in 1 ml BHI in vials containing a pre-measured amount of 500 µm garnet and 6mm zirconium grinding satellite beads as described above.

To determine if ingredients in cheese inhibited the RNA extraction or the PCR amplification of avian influenza virus (AIV), trials were conducted on cheese samples spiked with rgA/gyrfalcon/WA/41088/2014 PR8 low pathogenic avian influenza (LPAI) prior to liquification. Cheese was measured and added to BHI (w/ antibiotics) and spiked with 100µl of 10^3.5^ EID_50_/ml LPAI prior to homogenization. RNA extraction and PCR amplification were conducted as described below.

After liquification, both spiked and non-spiked samples were inoculated into eggs, as described below, to assess whether ingredients in dairy product samples would affect embryo mortality (non-spiked) or growth of AIV in eggs (spiked). Five eggs per sample were inoculated with 100 µl (each) homogenate and placed in an egg incubator at 37 ° C for 96 hours. Eggs were observed daily for signs of morbidity and mortality. At the end of 96 hours, allantoic fluid from all eggs was tested using the hemagglutination assay to determine the presence or absence of any viable hemagglutinating agent.

### Sample Processing

Samples were assigned unique identification numbers to blind the samples prior to shipping to the USDA ARS facility. Fluid milk and cream samples were aseptically portioned into 50 ml conical tubes prior to submitting the samples. Cheese, butter, cream cheese and ice cream samples were received without retail packaging aseptically repacked in Whirl-Pak bags. Liquid samples were placed at 4°C while solid samples were processed. Approximately 5–20 ml of ice cream was portioned into 50 ml conical tubes, while still frozen, upon arrival. These were placed at 4°C to allow them to melt prior to extraction while remaining portions were immediately frozen. Solid dairy samples (hard cheese, cream cheese, butter) were processed in 2-ml tubes, that contained a pre-measured amount of 500 micron garnet and 6mm zirconium grinding satellite beads (OPS Diagnostics, Lebanon, NJ). Approximately 250mg of each dairy solid sample was measured and added to 1 ml brain heart infusion broth and 30 µl antibiotics (initial concentration penicillin G, 10,000 IU/ml; gentamicin, 1 mg/ml; kanamycin, 0.65 mg/ml; streptomycin, 2 mg/ml; and amphotericin B, 20 µg/ml). Samples were liquified using a bead beater at 6.5 m/s for 60 seconds. The liquified sample was then used for RNA extraction similar to the liquid dairy samples.

### RNA Extraction

RNA was extracted from all samples using the MagMAX AI/ND Viral RNA extraction kit (Thermofisher Scientific, Waltham MA) according to manufacturer’s guidelines. Liquified solid dairy products and some ice cream samples required the use of 20-250 µl wide-opening pipet tips (Mettler Toledo Rainin, Columbus, OH) for adding samples to the lysis buffer. Extraction was completed using Kingfisher Duo (Thermofisher Scientific) benchtop machines. An internal positive control (VetMAX Xeno™ internal positive control-Liz assay kit, Thermofisher Scientific) was used during extraction to ensure components of samples did not interfere with the RNA extraction process. One microliter of a 10^-1^ dilution (final concentration: 1,000 copies) of control RNA was added to the MagMAX lysis buffer for each sample. Manufacturers guidelines recommend using 20,000 copies Xeno RNA per reaction, but in-house optimization determined 1,000 copies would give a detectable signal without interfering with amplification/signal detection for AIV matrix in the same reaction (data not shown). If both Xeno and matrix PCR were negative, samples were re-extracted using undiluted (10,000 copies) Xeno RNA.

### PCR

A quantitative real-time RT-PCR assay using primers and probe targeting the influenza A matrix gene was used to qualitatively detect AIV RNA in dairy samples. The 25 µl reaction mixture also included VetMAX primers and probe for the detection of the Xeno internal control and reagents from the Ag-Path ID RT-PCR kit: 12.5 µl Ag-Path RT PCR 2X buffer; 0.5 µl M +25 forward primer; 0.5 µl M-126 + 29nt reverse primer; 0.5 µl M+64 probe; 0.8 µl Xeno LIZ primer/probe mixture (provided in kit); 1.2 µl molecular-grade water; 1.0 µl Ag-Path enzyme; and 8 µl sample RNA. Samples were run on a Quantstudio 5 Real-time Detection System (Thermofisher Scientific) using the following cycling conditions: 1) reverse transcription at 45°C for 10 minutes 2) heat inactivation of RT and initial denaturation at 95°C for 10 minutes 3) 40 cycles of denaturation at 94°C for 10 seconds, annealing at 57°C for 30 s, and extension at 72°C for 10 seconds. The Quantstudio 5 spectral calibration did not include LIZ, so Cy5 (with similar emission/excitation) was used as an alternative channel to detect the internal positive control RNA. Non-infectious qRT-PCR-based quantity estimates were determined by comparing to a standard curve derived from RNA extracted from a 10-fold dilution series of quantified avian influenza virus stocks.(Spackman, 2020)

### Determination of Viability in Eggs

Samples positive for AIV RNA and samples that did not pass qRT-PCR extraction quality control were inoculated into 9-11 day-old embryonated eggs to determine viability of AIV in samples. For liquid samples, 30 µl of antibiotics (initial concentration penicillin G, 10,000 IU/ml; gentamicin, 1 mg/ml; kanamycin, 0.65 mg/ml; streptomycin, 2 mg/ml; and amphotericin B, 20 µg/ml) was added to 1ml of sample and incubated at room temperature for 30 minutes. Solid dairy homogenates (with previously added antibiotics) were removed from 4°C and were maintained at room temperature for 30 minutes. Liquified samples were spun briefly in a microcentrifuge to pellet beads and clarify the solution before inoculation using the supernatant. Five eggs were used for each sample with approximately 0.1 ml of sample/homogenate inoculated into each egg. Eggs were monitored for 96 hours and any deaths recorded daily. At the end of 96 hours, allantoic fluid from all eggs was tested using the hemagglutination assay to determine the presence or absence of any viable hemagglutinating agent.

### Nanopore Sequencing and Analysis

All qRT-PCR-positive samples were subject to the amplification of the hemagglutinin gene (HA) for further sequencing. The HA gene amplification was performed using the SuperScript IV One-Step RT-PCR kit (Thermofisher Scientific) in 50 μL reaction volumes comprised of 5 μL of total RNA, 25 μL of 2X Platinum SuperFi RT-PCR Master Mix, 2.5 μL of the forward primer H5+146EA: GTTACTGTTACACATGCCCA (10 pmol/μL) and 2.5 μL of the reverse primer H5-1347EA: AGTTCAGCATTATAAGTCCA (10 pmol/μL), 0.5 μL of SuperScript IV RT Mix, and 14.5 μL of sterile nuclease-free water. The test included an initial RT step for 10 min at 55°C, RT inactivation/initial denaturation for 2 min at 98°C, PCR steps of 40 cycles (10 seconds at 98°C, 10 seconds at 57.2°C, and 1 min at 72°C), and final extension for 5 min at 72°C. After thermocycling, 5 μL of the product was visualized on a 1.5% agarose gel to verify the amplification.

Nanopore sequencing libraries were prepared using the Native Barcoding Kit 24 V14 (SQK-NBD114.24, Oxford Nanopore Technologies). The final libraries were quantified using the High Sensitivity D5000 Screen Tape on a 4150 TapeStation (Agilent Technologies). We then sequenced a total of 29 samples in three batches using Flongle FLO-FLG114 and MinION FLO-MIN114 flow cells using the Mk1C sequencer with the MinKNOW 24.02.16 software in accordance with the manufacturer’s recommendations. Sequencing was run for 6 hours on Flongle and 3 hours on MinION flow cells.

The Nanopore raw Pod5 files were basecalled with a high-accuracy algorithm to generate FastQ files, which were then demultiplexed and trimmed using Dorado 7.3.11 within the MinKNOW 24.02.16 (bionic) software on a MinION Mk1C instrument. Short reads below 200 base pairs (bp) were removed during the sequencing run. Reads with a minimum quality of 9 were considered for further analysis. Consensus assembly of Nanopore reads was performed on the Galaxy platform.

Sequence data were assembled using a *de novo* approach utilizing Flye version 2.9.4 (Lin et al., 2016) . All final HA consensuses were called from the filtered reads aligned to *de novo* generated contigs using minimap2 (Li, 2018) and verified in Geneious Prime 2023.0.1. The coverage of the influenza virus genome was obtained using SAMtools depth(Danecek et al., 2021). Final consensus sequences were generated using the bam2consensus tool (Volkening, 2023) and polished using the medaka consensus pipeline (Ltd., O. N. T. (2020). *medaka*. https://github.com/nanoporetech/medaka). The viral sequences were submitted to GenBank.

### Phylogenetic Analysis

Phylogenetic analysis was conducted based on 1,171 nucleotide (nt) of HA gene. Forty-two available HA gene sequences from dairy cattle were retrieved from GISAID. Multiple sequence alignment was performed using MAFFT v7 (Katoh & Standley, 2013). Phylogenetic trees were constructed using the Maximum Likelihood method with general time reversible model using MEGA 7 (Kumar, Stecher, & Tamura, 2016). Evolutionary distances between sequences from this study were calculated based on a total of 1,171 nt positions using MEGA 7.

## Results

Different methods were evaluated to determine an effective way to both extract viral RNA from different types of cheeses and assure that the liquified cheese did not negatively affect virus culturing methods in embryonating chicken eggs (ECE). A variety of mechanical processes were compared to liquify or suspend the different cheeses so that it allowed efficient RNA extraction and virus culturing. Some test trials included spiked LPAI virus, rgA/gyrfalcon/WA/41088/2014, that was used to evaluate RNA extraction efficiency and evaluate growth in ECE. The four methods of liquifying by hand, grinding with a pestle in a 2ml vial, mincing with scalpel and vortexing, expelling through a 3-ml syringe (no needle), and pressing through a fine wire mesh filter did not liquify the sample enough to allow for virus culturing because it was too thick to pass through a 23 gauge needle. The mechanical extraction method using different types of lysis beads and a bead beating grinder and lysis system that is commonly used to extract RNA from animal tissue samples, provided adequate liquification of the sample. The 500 micron garnet and 6mm zirconium beads provided the best results for RNA extraction and was further tested for effects on virus culture in ECE.

The liquified cheese products were inoculated into 9–11 day old ECE and initially evaluated for toxicity of the embryos for 4 days of incubation. The cheese was either diluted at 1:2 or 1:4 (250m g cheese/0.5 or 1.0 ml of BHI) and inoculated into the ECE. The 1:2 dilution into eggs was not well tolerated with 7 of the 25 inoculated ECE having mortality after 24 hours in samples prepared with 5 different kinds of cheese. The cheese samples with the 1:4 (0.25 grams cheese/1.0 ml BHI) dilution of BHI was better tolerated with no mortality after 24 hours. Additional trials to assess virus culturing were all conducted with the 1:4 dilution. The cheese samples were spiked with LPAI virus and inoculated into ECE and incubated for 4 days. At the end of the 4 days, all the ECE were evaluated for hemagglutinating activity which if positive indicates live virus replication. Because cheeses are a fermented dairy product and each type of cheese uses unique bacterial or fungal cultures, a variety of cheeses were evaluated including cheddar, mozzarella, parmesan, American, 2 different types of blue cheese, gouda, and a raw summer milk cheese. All the virus-spiked cheese samples had an expected mortality pattern in the ECE on the third or fourth day after inoculation and positive hemagglutination of the allantoic fluid which indicates viral replication.

Using both the optimized protocol for solid dairy products and previously established methods for fluid dairy products, a total of 167 dairy samples were screened by real-time RT-PCR targeted to the influenza matrix gene with an internal positive control. The internal positive control, Xeno™, which is commercially available phage RNA that is added at the RNA extraction step, supported that almost every sample had good extraction efficiency of the phage RNA and was detected by the internal control real-time RT-PCR assay which also supports that no PCR inhibitors were present that affected the results. A total of 8 samples failed the internal positive control and were reextracted. A total of 4 samples failed the internal control standard on the second extraction, and these samples were inoculated into ECE to evaluate for viable virus, and were all negative. A total of 29 samples were positive for viral RNA from products processed in 7 different states (CO, ID, KY, MI, OH, SD, TX). The qRT-PCR confirmed products included fluid milk (7-ID, 1-KY), cheddar cheese (1-TX, 1-SD), 6 mozzarella (4-ID, 1-TX, 1-CO), 1 processed cheese (1-TX),3 butter (1-TX, 1-ID, 1-OH), and 9 ice cream (1-ID, 7-TX, 1-MI) samples (Table 1). The extrapolated titers of samples based on comparison to a standard curve show titers ranging from 1.2 to 4.7 log_10_ 50% egg infectious doses (EID_50_) per ml (Table 3). All qRT-PCR positive samples were inoculated into embryonating chicken eggs, and no viable virus was detected in any sample.

**Table 1.** Detection of Influenza A in retail dairy products by quantitative real-time RT-PCR.

| **Product** | **Number of Retail Product Samples** | **Retail Product Samples Positive for Viral RNA** | **Percent of Retail Product Samples Positive for Viral RNA** | **Detection of Live Virus in Retail Product** |
| --- | --- | --- | --- | --- |
| Skim Milk | 7 | 0 | 0.0% | No |
| 1% Milk | 11 | 2 | 18.2% | No |
| 2% Milk | 23 | 1 | 4.3% | No |
| Whole Milk | 38 | 5 | 13.2% | No |
| Heavy Cream | 1 | 0 | 0.0% | No |
| Cheddar Cheese | 14 | 2 | 14.3% | No |
| Mozzarella Cheese | 12 | 6 | 50.0% | No |
| Processed Cheese | 6 | 1 | 16.7% | No |
| Cream Cheese | 3 | 0 | 0.0% | No |
| Butter | 7 | 3 | 42.9% | No |
| Ice Cream | 22 | 9 | 40.9% | No |
| Aged Raw Milk Cheese | 23 | 0 | 0.0% | No |
| **Total** | **167** | **29** | **17.4%** | **None** |

**Table 2.**
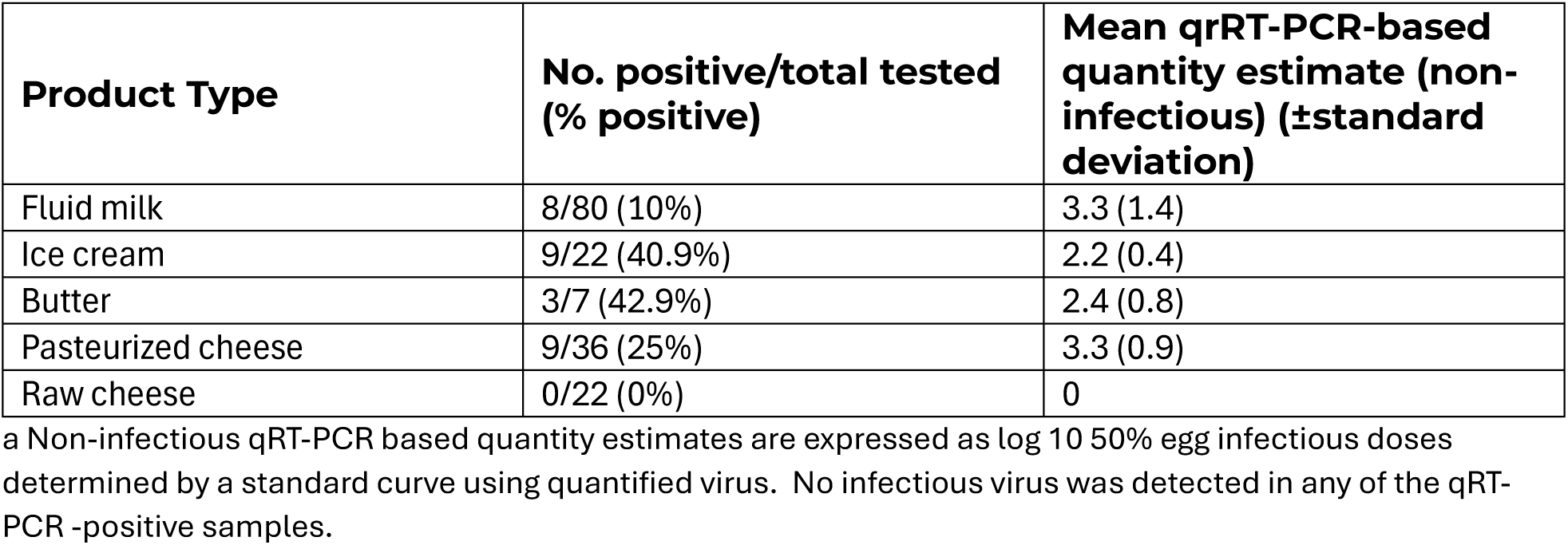
Extrapolated viral titers from dairy samples by real-time RT-PCR^a^.

| **Product Type** | **No. positive/total tested (% positive)** | **Mean qRT-PCR-based quantity estimate (non-infectious) (±standard deviation)** |
| --- | --- | --- |
| Fluid milk | 8/80 (10%) | 3.3 (1.4) |
| Ice cream | 9/22 (40.9%) | 2.2 (0.4) |
| Butter | 3/7 (42.9%) | 2.4 (0.8) |
| Pasteurized cheese | 9/36 (25%) | 3.3 (0.9) |
| Raw cheese | 0/22 (0%) | 0 |

<sup>a</sup> Non-infectious qRT-PCR based quantity estimates are expressed as log<sub>10</sub> 50% egg infectious doses determined by a standard curve using quantified virus. No infectious virus was detected in any of the qRT-PCR -positive samples.

**Table 3.** Summary statistics of HA gene sequencing run.

| **Sample No.** | **Sample Type** | **qRT-PCR, Ct** | **Flow cell** | **AIV reads** | **Mean Depth** | **GenBank Acc. No.** |
| --- | --- | --- | --- | --- | --- | --- |
| 1 | Mozzarella | 31.1 | MinION | 40,021 | 39,752 | PQ162248 |
| 2 | Mozzarella | 31.4 | MinION | 606 | 525 | PQ162249 |
| 3 | Cheddar | 26.5 | MinION | 20,989 | 20,844 | PQ162250 |
| 4 | Cheddar | 24.7 | MinION | 50,340 | 50,121 | PQ162251 |
| 5 | ice cream | 27.9 | MinION | 6,688 | 6,588 | PQ162252 |
| 6 | whole milk | 35.2 | MinION | 318 | 267 | PQ162253 |
| 7 | whole milk | 22.9 | MinION | 63,345 | 63,406 | PQ162254 |
| 8 | whole milk | 23.3 | MinION | 76,444 | 76,199 | PQ162255 |
| 9 | Mozzarella | 26.9 | MinION | 49,908 | 49,584 | PQ162256 |
| 10 | Mozzarella | 26.3 | MinION | 39,108 | 38,994 | PQ162257 |
| 11 | low fat milk | 28.6 | MinION | 23,929 | 23,772 | PQ162258 |
| 12 | whole milk | 30.0 | MinION | 8,719 | 8,629 | PQ162259 |
| 13 | whole milk | 29.6 | MinION | 92,783 | 27,294 | PQ162260 |
| 14 | 2% milk | 29.8 | MinION | 21,139 | 20,975 | PQ162261 |
| 15 | Mozzarella | 27.0 | MinION | 22,964 | 22,733 | PQ162262 |
| 16 | Mozzarella | 25.9 | MinION | 18,815 | 18,630 | PQ162263 |
| 17 | whole milk | 24.5 | Flongle | 432 | 420 | PQ162264 |
| 18 | fat free milk | 23.8 | Flongle | 612 | 605 | PQ162265 |
| 19 | 1% low fat milk | 26.6 | Flongle | 320 | 314 | PQ162266 |
| 20 | whole milk | 27.2 | Flongle | 3,573 | 3,542 | PQ162267 |
| 21 | 2% reduced fat milk | 24.3 | Flongle | 6,827 | 6,761 | PQ162268 |
| 22 | vitamin d milk | 24.3 | Flongle | 12,684 | 12,528 | PQ162269 |
| 23 | whole milk | 26 | Flongle | 16,080 | 15,949 | PQ162270 |
| 24 | fat free milk | 25.9 | Flongle | 19,748 | 19,437 | PQ162271 |
| 25 | 1% low fat milk | 23.4 | Flongle | 6,413 | 6,364 | PQ162272 |
| 26 | 2% reduced fat milk | 23.1 | Flongle | 11,993 | 11,785 | PQ162273 |
| 27 | vitamin d milk | 24 | Flongle | 3,883 | 3,801 | PQ162274 |
| 28 | 2% reduced fat milk | 21 | Flongle | 17,387 | 17,118 | PQ162275 |
| 29 | 2% reduced fat milk | 24.6 | Flongle | 4,333 | 4,242 | PQ162276 |

To further characterize the viral RNA, the H5 hemagglutinin gene was PCR amplified and the amplicons were sequenced on the MK1C sequencer and analyzed. The PCR amplicon size of 1,171 bp, which is approximately 2/3rds of the hemagglutinin gene was amplified and compared to available avian influenza sequences. The HA gene amplification was not successful on all the qRT-PCR matrix positive samples (16 out of 29). Samples from this study were sequenced on a MinION flow cell. An additional 13 samples, which included the earlier retail dairy milk positive samples, were sequenced on Flongle flow cells(Spackman et al., 2024). All sequenced samples had acceptable sequence read number and depth of min 50X (Table 3).

Obtained nucleotide sequences of amplified samples were highly similar with *p*-distance ranging from 0.000 to 0.005. Amino acid changes were observed only in 6 samples: cheddar (n = 1), low fat milk (n = 1), 1% low fat milk (n = 2), 2% milk (n = 1), and whole milk (n = 1).Sequences from retail dairy samples from this study were also highly similar to the previously reported HPAI viruses detected in dairy cattle, which provides evidence that the viral RNA detected is from a single source introduction into dairy cattle (Figure 1).

**Figure 1.**
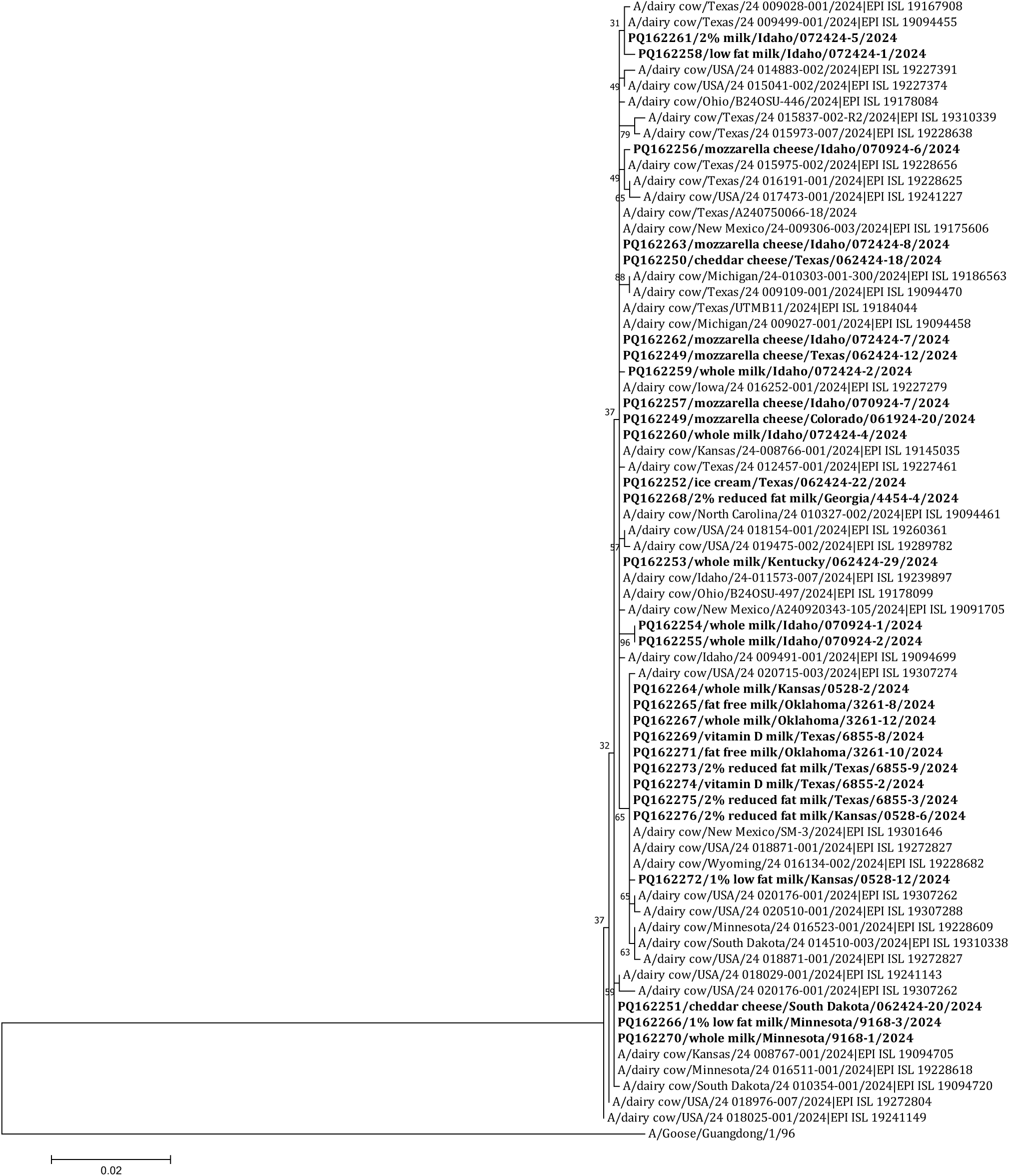
Phylogenetic analysis of hemagglutinin gene sequences of highly pathogenic avian influenza A (H5N1) viruses from US dairy products (highlighted in bold) together with viruses from US dairy cattle. The tree is drawn to scale, with branch lengths measured in the number of substitutions per site. The analysis involved 72 nucleotide sequences. There were a total of 1168 positions in the final dataset. Evolutionary analyses were conducted in MEGA7.

## Discussion

This study was an extension of an earlier retail dairy milk survey by increasing the geographic collection of samples and evaluating different types of dairy products that were not surveyed in the first study including several different types of cheeses, ice cream and butter. A total of 17.4% of all the products tested had detectable viral RNA, but all samples were negative for viable virus as assessed through culture in ECE. These results support the first study that found similar results. In the first study, all the samples evaluated were products that are required to be pasteurized as required by FDA regulation for interstate commerce, and high temperature/short time continuous flow pasteurization was recently shown to complete inactivate HPAI virus in milk (Spackman submitted). Therefore, both experimental evaluation and the first dairy survey support that current FDA pasteurization requirements provide broad protection for consumers for most dairy products. This study however included 23 cheese samples that used unpasteurized or raw milk for cheese production in accordance with FDA requirements for aged raw milk cheeses. The production of raw milk cheese for interstate commerce is also regulated by the FDA for interstate commerce and requires that raw milk cheeses be aged for a minimum of 60 days at no lower than 35°F (21 CFR Part 133). Aged raw milk cheeses are the only dairy product for human consumption produced with unpasteurized milk that can be sold in interstate commerce. Although 23 aged raw milk cheeses were included in this survey, all samples were negative for the detection of viral RNA which suggests that the milk used to prepare these cheeses were from uninfected cows. Because there was no evidence of virus in the cheese, we can’t draw any conclusions on whether the current requirements of 60 days of aging are sufficient to inactivate viable virus.

It should also be recognized that cheese production, for both pasteurized and aged raw milk cheeses, is not a homogeneous process and cheeses are typically heated to different temperatures during processing, different bacterial or fungal cultures are added to provide the unique characteristics of different types of cheese, cheeses achieve different levels of acidic pH, and different cheeses are aged for different periods of time. Each of these steps in the cheese making process could play an impactful role in the inactivation of pathogens including HPAI. For aged raw milk cheeses, the 60-day minimum aging requirement might eliminate or reduce potential pathogen levels for the safety of the consumer. However, the HPAI outbreak in dairy cows presents a unique threat to consumers because it is a potential zoonotic pathogen, and it is unclear if the procedures that are or might be protective against bacterial pathogens are also effective for viral pathogens. It should be noted that AIVs are as or more sensitive to thermal inactivation as bacterial pathogens like Salmonella, and that cooking temperatures that kill Salmonella sp. will also inactive influenza viruses(Chmielewski, Beck, & Swayne, 2011, 2013; Luchansky et al., 2024; Thomas & Swayne, 2007, 2009). Probably the most studied viral pathogen in milk from dairy cattle is foot and mouth disease (FMD) virus, and this virus was studied because of concerns about transmission of this virus from endemic countries to cattle in FMD free countries through the importation of dairy products. A single study examined the virus recovery in cheeses made from FMD contaminated milk and found that cheese made from heated milk was negative for virus recovery. However, cheddar cheese made from raw milk was positive for virus after 60 days of aging, but negative after 120 days of aging. Viable virus was not detected in mozzarella cheese and it was not found in Camembert cheese at 35 days of aging(Blackwell, 1976). It should be understood that FMD is more heat and environmentally resistant than HPAI virus and may not be the best surrogate for comparison. The FMD cheese study does clearly show that different cheeses, which are prepared in different ways, may have virus inactivated during the heating steps, direct degradation by the bacterial or fungi used for cheese making, or potentially the different pH levels that are achieved in the cheese making process. Further studies are planned with raw milk cheeses to evaluate if current aging requirements are sufficient to inactivate HPAI virus.

Another goal of this research study was to develop and validate procedures for sampling solid dairy products to detect viral RNA and assure culturing techniques were adequate. The previous retail milk survey documented that the RNA extraction methods and culturing techniques for liquid milk products, with minor modifications, were effective at detecting viral RNA and other studies had shown that virus could easily be recovered in liquid milk samples. For culturing in ECE, higher levels of antibiotics an antimycotics were included to prevent bacterial or fungal infections in eggs(Spackman et al., 2024). The current retail surveys included cheese, butter and ice cream that had not been tested previously. The ice cream after melting was handled similarly to liquid milk, but the cheese and butter required additional processing steps. After careful optimization, a modified procedure that is used with avian tissue samples was adopted to use grinding beads in a bead beater to liquify the sample(Spackman, 2020). More BHI media was used to dilute the sample because of the viscosity of the sample, but RNA extraction worked both for retail cheese and butter samples as demonstrated with products spiked with avian influenza virus and detection of the internal positive control. The second concern was the potential for the cheese or residual bacteria or fungal culture in the cheese to be toxic to the ECE which would invalidate the culture process or inhibit the viral replication in ECE. Using spiked in virus with different cheeses, we did not detect any adverse effects on the ECE and viable virus could be detected from the inoculated ECE. One alternative method using Trizol and proteinase K for extraction of Norovirus RNA was also recently reported and could provide an alternative approach(Cantelli et al., 2024), but this approach was not evaluated in this study.

The sequence analysis of the samples from both this survey and the previous retail milk survey showed that there is likely little antigenic variation in the hemagglutinin protein. This lack of antigenic variation supports a single introduction of the B3.13 variant into dairy cattle and its subsequent spread within the dairy industry. The first retail dairy survey also tested samples by a H5 real-time RT-PCR that confirmed all the samples were HPAI viruses of the 2.3.4.4 lineage.

However, when sequencing, despite the samples testing matrix positive, a percentage of them failed to produce quality H5 sequence that could be analyzed. There are two likely reasons for the sequence failures including that there was not enough viral RNA to sequence the larger genomic amplicon or that the viral RNA was fragmented which prevented amplification of larger amplicons. Many of the matrix positive samples, which targets an amplicon slightly over 100bp, had cycle threshold (Ct) values indicating only low levels of detected virus. In general, the larger the amplicon, the less efficient the PCR amplification which means many of the samples that had low levels of viral RNA likely were sequence negative because of this decreased PCR efficiency because for sequencing a much larger target, 1171 bp was targeted. The second possibility is that the homogenization and pasteurization of the different milk products, which effectively inactivates the viral RNA, also negatively effects RNA quality and fragments the viral RNA. The potential for fragmentation is supported by an earlier unsuccessful attempt (data not shown) of whole genome viral amplification and sequencing which uses the highly conserved terminal sequences of influenza as primer sequences. This whole genome amplification approach is the most commonly used method for influenza sequencing and is used both pure culture and influenza in clinical samples (Hoffmann et al., 2001; Wang et al., 2015). The complete failure of this sequence attempt supports the idea of fragmentation of the viral RNA. Additionally, some samples with higher viral RNA levels could not be sequenced, so viral RNA levels were not always a good predictive of successful amplification. Alternative approaches for amplification can potentially allow full genome sequencing but are beyond the scope of the report.

This retail dairy survey provides additional support for the robustness of the established pasteurization program for dairy products. The revised methodology allows for the detection of viral RNA in cheese, butter, and ice cream samples, which had not previously been tested. Because most cheese in the U.S. is produced from pasteurized milk, the risk of live virus in these products was already extremely low. In addition, further processing and the microbial fermentation used in cheese are likely to have additional anti-viral activity. The negative virus isolation results should therefore have been the expectation for this study. Although the 17% positive detection rate of viral RNA appears to implicate a large number of dairies as having infected cows. This type of survey should not be used to estimate the number of farms with infected cows because the milk from retail samples represents an aggregate of many cows from many dairies, and the survey was not completely random in the selection of samples to test. Retail milk surveys can be used to provide additional consumer assurance of the safety of dairy products. The only samples tested from unpasteurized milk were the aged raw milk cheeses, and all those samples were negative.

Additional studies are needed to evaluate aged raw milk cheeses, but again the microbial fermentation with the reduced pH levels and the 60-day required aging will likely inactivate avian influenza in these products(Bansal & Veena, 2024; Hamilton, Whittaker, & Daniel, 2012; Russell, 2021). The risk of H5N1 HPAI infection to humans through drinking unpasteurized milk remains unknown at this time, but the recommendation to only drink pasteurized milk continues to be a prudent public health recommendation.

## Data Availability

All data produced in the present work are contained in the manuscript or is available in Genbank (sequence data)

## Acknowledgements

The authors gratefully thank, Suzanne DeBlois, and Ricky Zoller for technical assistance with this work, and Craig Kellogg and Brenda Kellogg for donations in support of the research.

We gratefully acknowledge authors from originating and submitting laboratories of the sequences from the GISAID databases. A table of the contributors is available in Supplementary Table S1

Any use of trade, firm, or product names is for descriptive purposes only and does not imply endorsement by the US Government. This research was supported by US Department of Agriculture (USDA)-Agricultural Research Service Project No. 6040-32000-081-00D and the US Food and Drug Administration InterAgency Agreement 6040-32000-081-037I. All opinions expressed in this paper are the authors’ and do not necessarily reflect the policies and views of the USDA or FDA. The USDA is an equal opportunity provider and employer.

